# Traumatic Amputations - A Nationwide Epidemiological Analysis of a developing country over 16 years

**DOI:** 10.1101/2024.09.05.24313153

**Authors:** Marcella Moura Ceratti, Carolina Carvalho Jansen Sorbello, Isabela Roskamp Sunye, Felipe Soares Portela, Marcelo Fiorelli Alexandrino da Silva, Marcelo Passos Teivelis, Miguel Cendoroglo Neto, Nelson Wolosker

## Abstract

**Background:** Defining the impact of amputation is essential for developing cost-effective preventive health policies. Trauma is one of the most common causes of limb loss, affecting mainly the young working population. To date, few studies have investigated the epidemiology of patients undergoing trauma-related amputations and their public health implications in developing countries. The aim of this study was to analyze all limb amputations due to traumatic injuries performed in the Brazilian public health system over a 16-year period, studying their incidence, demographics, hospitalization and costs.

**Objective:** To analyze the epidemiologic data on traumatic amputations within Brazil’s public health system.

**Methods:** This study was a cross-sectional and retrospective population-based analysis of traumatic amputations performed in the Brazilian public health system from 2008 to 2023. DATASUS, a public database of the Brazilian public health system, was used to select trauma cases, which were filtered by the traumatic amputation code. The dataset included the number of procedures, regional distribution, patient demographics, length of hospital stay, ICU stay, lethality and financial reimbursement.

**Results:** There were 202,940 traumatic amputations in Brazil between 2008 and 2023. This condition was most common in males (78.7%), with an average age of 44.77 years, mainly involving fingers (62.7%). Cases involving lower limbs led to longer hospital stays and more ICU admissions. The mean length of hospital stay was 4.57 days, culminating in a lethality of 3.15%, which remained stable during the follow-up period. The estimated total cost to the Brazilian public health system was US$ 54,870,097.79, equivalent to approximately US$ 34,29381 spent per year on traumatic amputations, with the lower limb representing twice as much per patient.

**Conclusion:** Traumatic amputation is still common in all regions of Brazil and has remained stable over the past 16 years, as has its lethality. Lesions leading to lower limb amputation account for the majority of hospital stays, intensive care unit admissions, and higher lethality and costs.

## INTRODUCTION

Traumatic amputations, characterized by the abrupt loss of a limb due to external forces, represent a significant medical challenge with varied incidence rates across different regions and contexts. A study conducted at a Level I trauma center in North India reported a 2.5% incidence among trauma cases (1). In contrast, in the United States, the incidence of major lower extremity amputations resulting from trauma was 0.18% of all trauma admissions (2).

In the United States, trauma is the second most common cause of limb loss, following vascular disease (3). The incidence and prevalence of traumatic amputations rose by 16.4% and 49.2%, respectively, from 1990 to 2019. However, age-standardized incidence and prevalence rates have declined over the years (4). In Brazil, a study on lower limb amputations showed that trauma is decreasing as a cause of amputation in the country, decreasing from 12% of amputations in 2008 to 7% in 2020. At the same time, diabetes became the leading cause of amputations (27%), followed by gangrene (25%) and peripheral arterial disease (23%) (5).

The causes of traumatic amputations vary depending on the location and time period. During wars, for example, the primary causes involve improvised explosive devices, as seen in conflicts in Afghanistan and Iraq. However, in the civilian population on a global scale, the leading cause of traumatic amputation before the age of 60 is exposure to mechanical forces. In contrast, after 60, the main cause is falls (4).

The lethality for traumatic amputations ranges from 6% to 13% in the literature (1,2). However, another significant problem with amputations in general is their morbidity and loss of functionality. Despite advances in adaptation techniques for individuals with amputated limbs, this type of injury continues to have a substantial impact on quality of life. The main implications of living with an amputated limb include complications of the surgical wound (such as infection and necrosis (6); pain (such as phantom limb pain in 10.9% to 50.9% of cases) (7); fractures due to osteoporosis and muscle atrophy (8); psychosocial distress and dependence on daily activities (9).

Although there are numerous studies analyzing the prevalence of amputations worldwide, no national epidemiological study has evaluated traumatic amputations in Brazil. Brazil is the largest country in Latin America with a population of 203,080,756 inhabitants (10), 71% of whom depend entirely on the Brazilian public health system (SUS - ‘Sistema Único de Saúde’) for healthcare services (11). All the information on SUS hospitalizations is available on a publicly accessible platform on the DATASUS (Departamento de Informação e Informática do SUS) website (12).

This study aimed to present the main epidemiological data on traumatic amputations in Brazil. Data from DATASUS, referring to the entire national territory, contain information on the number of traumatic amputations, body segment amputated, age, biological sex, lethality, hospital stay, need for an intensive care unit (ICU) and costs for the public health system.

## METHODS

### Study Design and Ethical Approval

This is a cross-sectional and retrospective population-based study. The institution’s Research Ethics Committee approved it (CAAAE 35826320.2.0000.0071). Since DATASUS is a public repository of anonymized data, informed consent was not required (12).

### Data Source, Extraction Process, and Period

Data were retrieved from DATASUS, the online database of the Brazilian Health System (SUS), which compiles information on government-funded hospitalizations nationwide. The focus was on trauma amputation procedures from 2008 to 2023. The extraction of data was automated using a Python-based protocol (v. 2.7.13; Beaverton, OR, USA) developed by our IT department, running under Windows 10. Selenium WebDriver (v. 3.1.8; Selenium HQ) and Pandas (v. 2.7.13; Lambda Foundry, Inc. and PyData Development Team, NY, USA) tools were used for data segregation and refinement.

### Study Population

The average Brazilian population from 2008 to 2022 was 196,918,278 (10). Considering that 71% of the population uses exclusively the Brazilian public health service, this study’s population sample was approximately 140,000,000 patients (11).

### Selection Criteria and Extracted Data

Data related to trauma cases were identified using ICD-10 codes (S00-S99 and T00-T98). From these, cases involving “disarticulation of the shoulder joint (scapulohumeral joint)”, “amputation/disarticulation of the hand and wrist”, “amputation/disarticulation of the upper limbs”, “reimplantation from the shoulder to the mid-third of the forearm”, “reimplantation from the distal third of the forearm to the metacarpals”, “reimplantation or revascularization at the level of the hand and other fingers (except the thumb)”, “reimplantation or revascularization of the thumb”, “hip disarticulation (coxofemoral disarticulation)”, “amputation/disarticulation of the lower limbs”, “amputation/disarticulation of the foot and tarsus”, “reimplantation from the thigh to the proximal third of the leg”, “reimplantation from the mid-third of the leg to the foot”, and “amputation/disarticulation of a finger”. The corresponding DATASUS codes are, respectively: 0408010070, 0408020016, 0408020024,0408020253,040802026, 04080202701, 0408020288, 0408040106, 0408050012, 0408050020, 0408050306, 0408050314, 0408060042 . The dataset included annual procedure counts, regional distributions, patient demographics (age and biological sex), lengths of hospital stays, ICU durations, lethality and financial reimbursements. Financial data in Brazilian Reais (R$) was converted to U.S. dollars (U$), based on the median exchange rate from 2008 to 2023, amounting to U$ 1 = R$ 3,2018 (13).

### Data Preparation

Data compilation and tabulation were performed in .csv format using Microsoft Office Excel 2019 (Redmond, WA, USA). Population data by age group were obtained from the Brazilian Institute of Geography and Statistics (IBGE) (14). Statistical analyses were based on the population relying solely on the Brazilian public health system, which is approximately 140,000,000 patients (11).

### Statistical Analysis

Statistical analyses were conducted using SPSS version 20.0 for Windows (IBM Corp, Armonk, NY). Linear regression was employed to analyze trends in traumatic amputations across various age groups. Chi-square and likelihood ratio tests were used to evaluate differences in trauma rates and lethality across the country’s macro-regions. A significance level of P ≤ 0.05 was set for all statistical tests.

## RESULTS

From 2008 to 2023, there were 202,940 traumatic amputations. Among these, the majority (62.7%) were finger amputations, 29.6% lower limb amputations, and 7.7% upper limb amputations. Over the years of study, there was an increase in the incidence of finger amputations at a rate of 1.4% per year (incidence ratio per year = 1.014 [95% CI: 1.011, 1.018]; p-value < 0.001). In contrast, the incidence of upper and lower limb amputations remained unchanged, showing no statistically significant differences (incidence rate of lower limb amputations per year = 0.992 [95% CI: 0.984, 1.001]; p-value = 0.073, and 1.000 [95% CI: 0.988, 1.013]; p-value = 0.939, respectively). **Figure 1** shows the number of traumatic amputations over the year.

**Figure 1.**
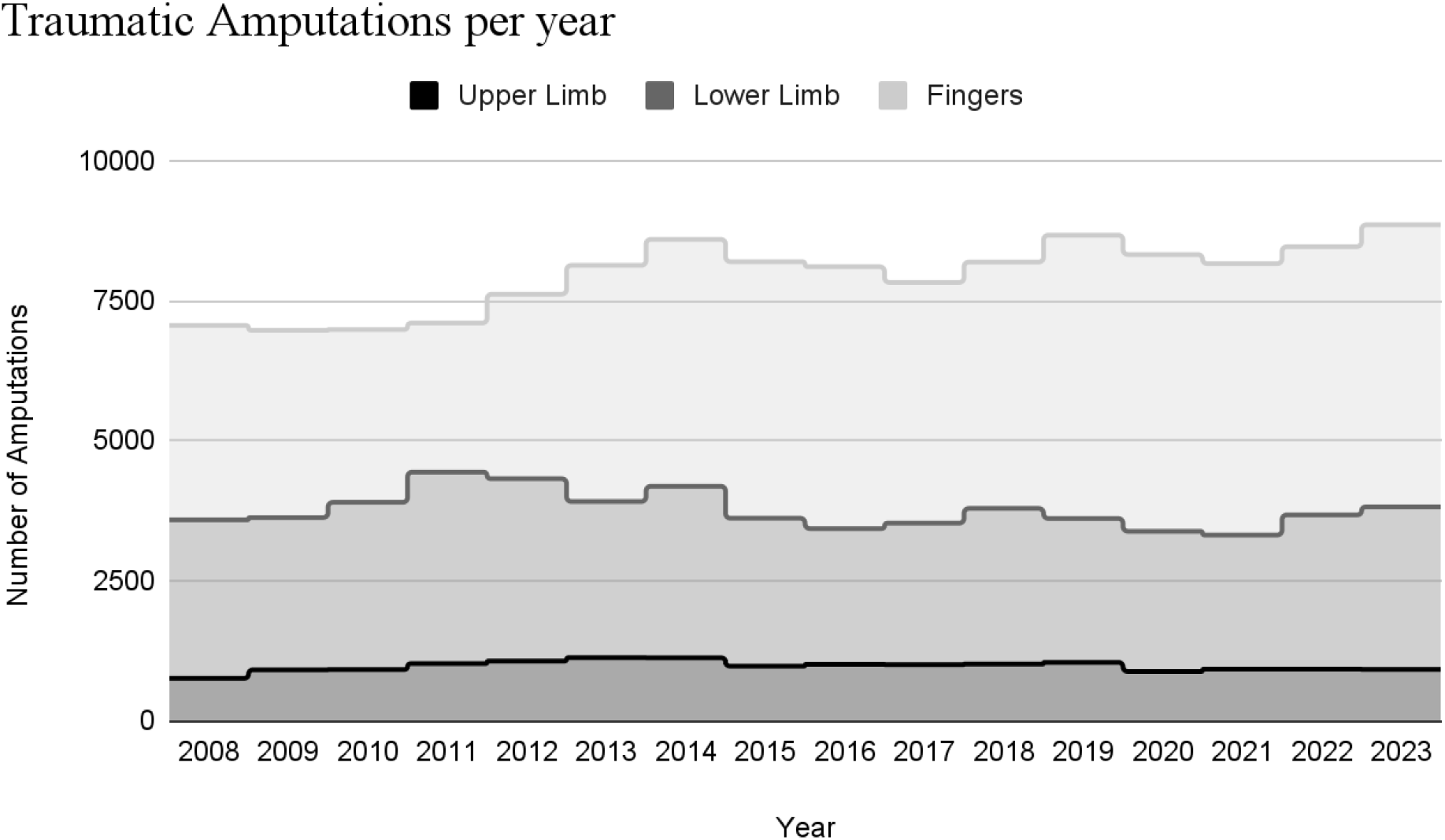
Number of traumatic amputations from 2008 to 2023.

**Figure 2** shows the distribution of traumatic amputations across the five regions of Brazil. Most amputations occurred in the Northeast and Southeast regions. Adjusting for population size (10), the region with the highest incidence of traumatic amputations over 16 years was the south (122 amputations per 100,000 inhabitants), followed by the northeast (119 per 100,000 inhabitants), the center-west (116 per 100,000 inhabitants), the north (101 per 100,000 inhabitants) and finally the southeast (77 per 100,000 inhabitants).

**Figure 2:**
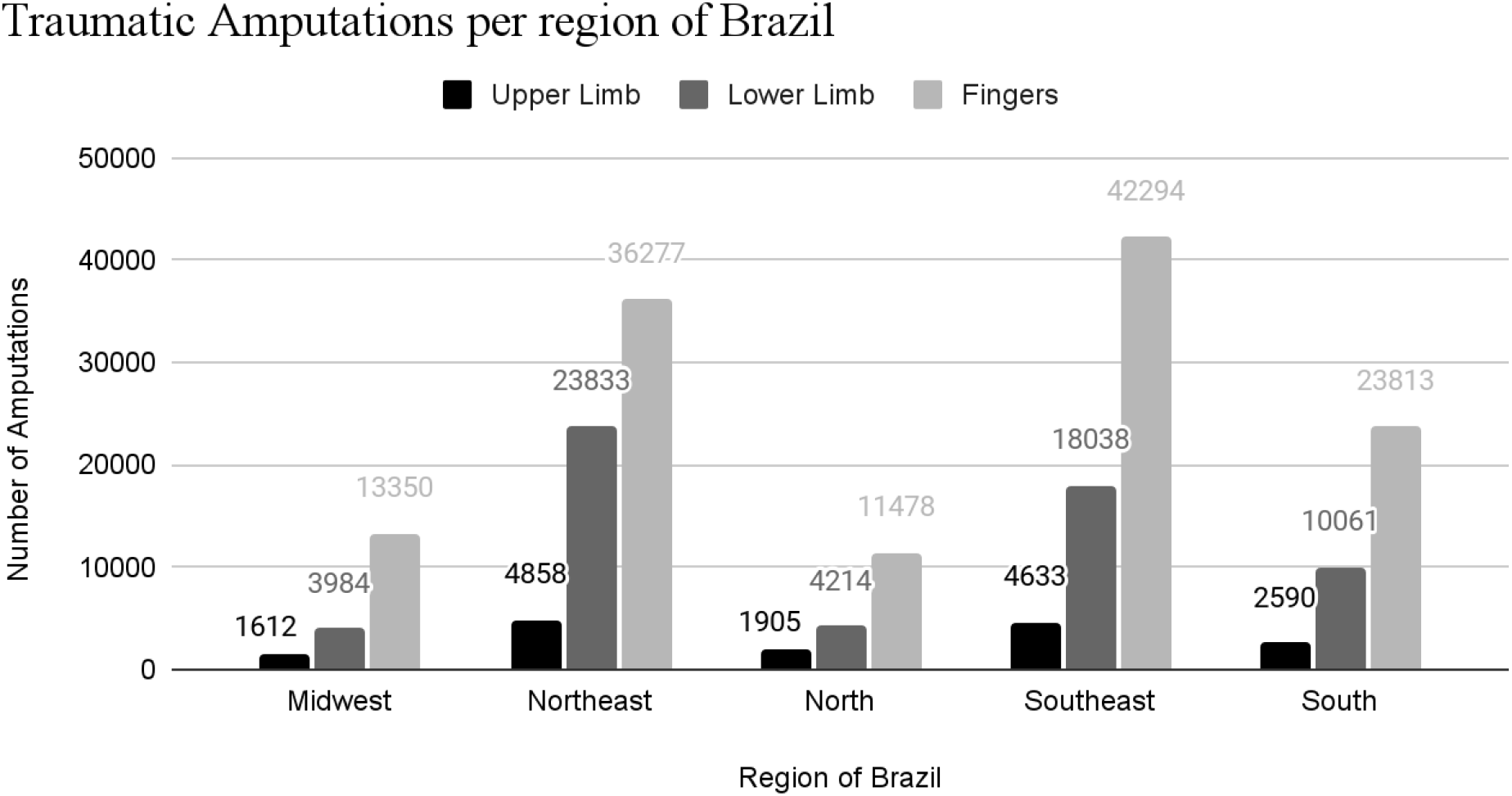
Number of Traumatic Amputations per region of Brazil.

Traumatic amputations were more common in males (78.7%). For finger amputations, males accounted for 82.6%; for upper limb amputations, 84.1%; and for lower limb amputations, 69% (p-value < 0.001).

Traumatic amputations were more common between the ages of 20 and 59 (**Figure 3**). The mean age was 44.77 years. Cases of lower limb amputations peaked around the ages of 65-69 and again in patients aged 80 or over. In the pediatric age group, the vast majority of amputations were due to finger amputations.

**Figure 3.**
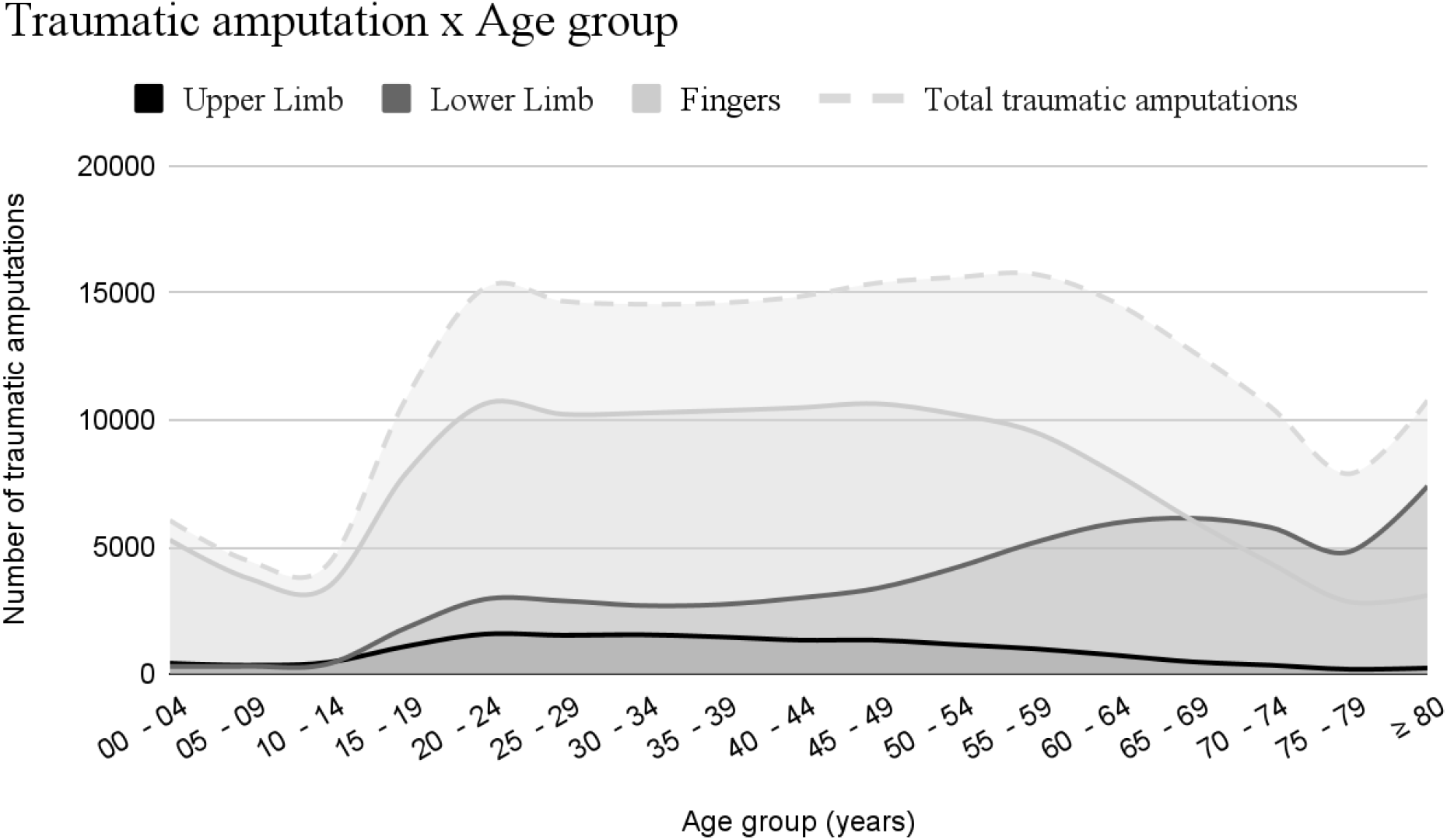
Number of patients with traumatic amputations per age group.

The majority of lower limb amputations (40.2%) occurred in the age group of 65 years or older; however, only 12.9% of finger amputations and 8.6% of upper limb amputations occurred in this age group (p-value < 0.001). Furthermore, 9.8% of finger amputations and 8.3% of upper limb amputations occurred in the pediatric age group, while only 1.8% of lower limb amputations occurred in this age group (p-value < 0.001).

**Figure 4** shows the number of hospitalization days for traumatic amputations of lower limbs, upper limbs, and fingers. Most patients stayed in the hospital for one day. The mean duration of hospitalization was 4.57 days. As expected, patients with lower limb amputations stayed in the hospital for a longer duration, with most being hospitalized for 3 days.

**Figure 4:**
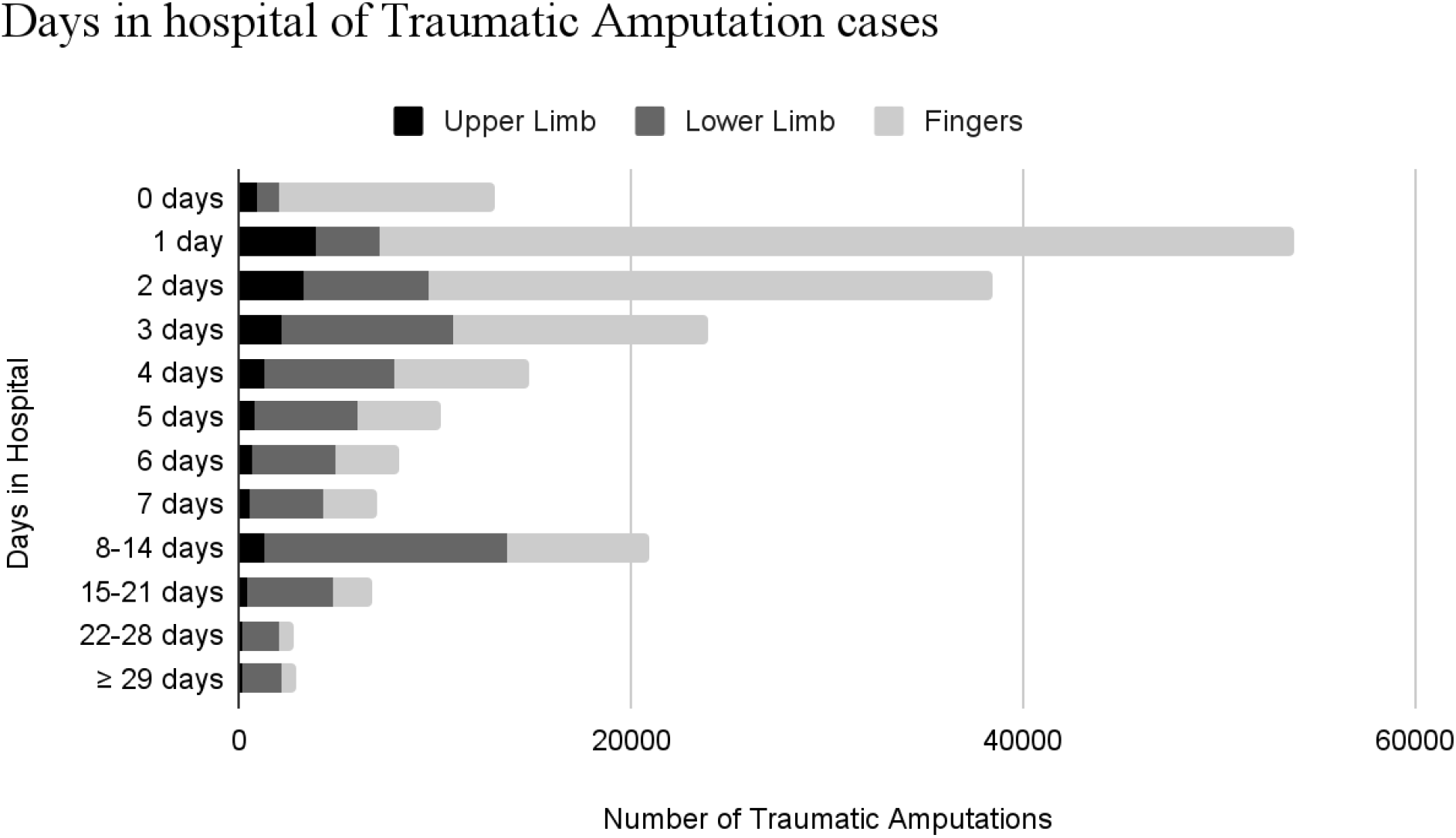
Days in hospital of patients with traumatic amputations.

**Figure 5:**
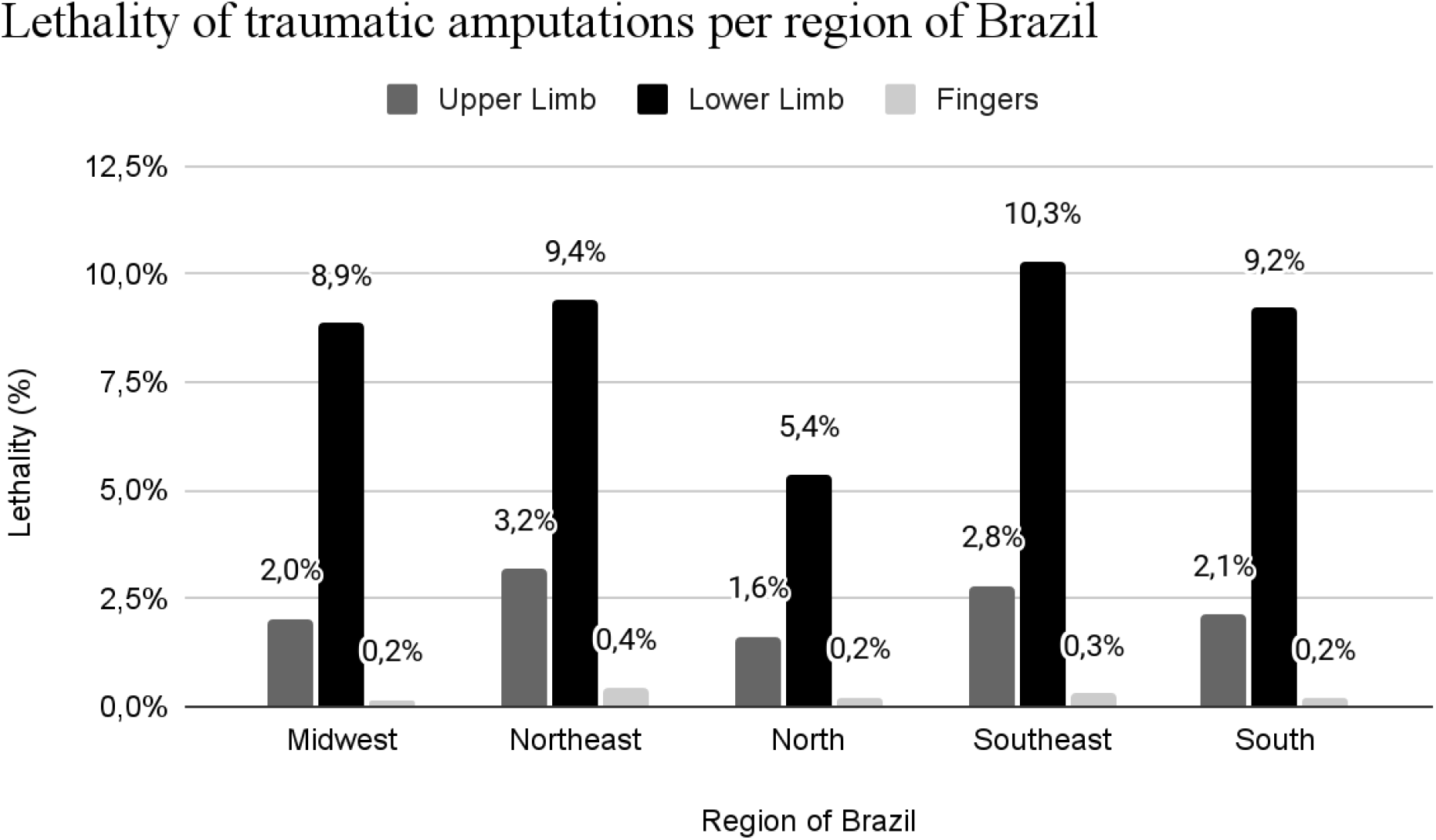
Lethality of traumatic amputations per region of Brazil. (p-value < 0.001).

The majority of patients with finger amputations (67.9%) and upper limb amputations (51.9%) required hospitalization for 0 to 2 days, while the majority of lower limb amputations were hospitalized for 3 to 7 days (p-value < 0.001). 13.6% of patients with lower limb amputations required hospitalization for 15 days or more, compared to only 2.7% of patients with finger amputations and 5.2% of patients with upper limb amputations (p-value < 0.001).

The vast majority of patients with traumatic amputations (94%) did not require intensive care. For those who needed ICU care, the average length of stay in the ICU was 5.85 days. Patients with lower limb amputations required more hospitalization in the intensive care unit (ICU), with 17% of them being admitted to the ICU, compared to 8.1% of patients with upper limb amputations and 0.6% of those with finger amputations (p-value < 0.001) as shown in **Table 1**.

**Table 1.** Days in Intensive Care Unit (ICU) for patients with traumatic amputations.

|  | Traumatic Amputations |  |  | p-value |
| --- | --- | --- | --- | --- |
|  | Fingers | Lower Limb | Upper Limb |  |
| Days in ICU |  |  |  |  |
| 0 | 126456 (99.4) | 49908 (83.0) | 14340 (91.9) | <0.001 |
| 1 - 5 | 495 (0.4) | 6736 (11.2) | 830 (5.3) |  |
| 6 or more | 261 (0.2) | 3486 (5.8) | 428 (2.7) |  |
|  | 127212 | 60130 | 15598 |  |
| Total | (100.0) | (100.0) | (100.0) |  |
| Need for ICU |  |  |  |  |
| No | 126456 (99.4) | 49908 (83.0) | 14340 (91.9) | <0.001 |
| Yes | 756 (0.6) | 10222 (17.0) | 1258 (8.1) |  |
|  | 127212 | 60130 | 15598 |  |
| Total | (100.0) | (100.0) | (100.0) |  |

The overall lethality of traumatic amputations was 3.15%.The highest lethality was in the lower limb amputation group (9.32%), followed by upper limbs (2.59%) and fingers (0.30%), with a statistically significant difference (p-value < 0.001) . The region with the highest lethality was the Southeast, followed by the Northeast (**Figure 6**).

**Figure 6:**
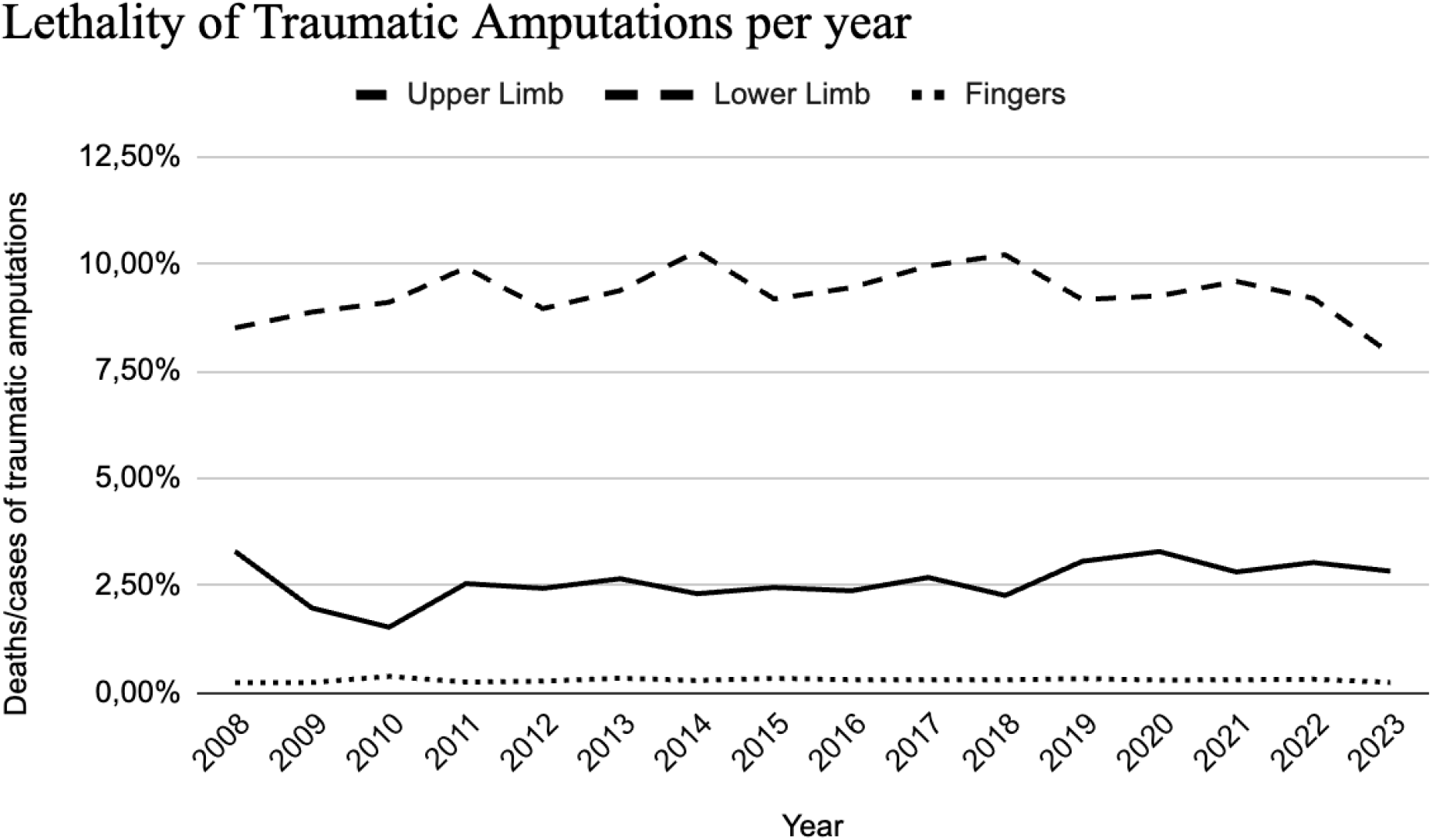
Lethality of traumatic amputations per year.

Lethality of traumatic amputations remained stable from 2008 to 2023, with no statistically significant variations for traumatic finger amputations (lethality ratio per year = 1.004 [95% CI: 0.986, 1.023], p-value = 0.632), upper limb amputations (lethality ratio per year = 1.017 [95% CI: 0.994, 1.041], p-value = 0.146), and lower limb amputations (lethality ratio per year = 1.000 [95% CI: 0.991, 1.009], p-value = 0.986). The lethality over the years can be observed in **Figure 7**.

The Brazilian public health system spent a total of $54,870,097.79 on traumatic amputations from 2008 to 2023. Most of the expenses occurred in the Southeast and Northeast regions, which is consistent with the number of cases in these areas. Additionally, the treatment of traumatic lower limb amputations was more costly than other types of amputations. **Table 2** provides more details. The overall cost per patient for traumatic amputations was $270.38. For lower limb amputations, this cost increased to $542.81.

**Table 2.** Cost of treatment for traumatic Amputations by Region in Brazil.

|  | Total Cost<br>(american dollars) | Mean Cost per patient<br>(american dollars) |
| --- | --- | --- |
| **Fingers** |  |  |
| Midwest | 1954574.99 | 146.41 |
| Northeast | 5253346.35 | 144.81 |
| North | 1663599.75 | 144.94 |
| Southeast | 6303489.39 | 149.04 |
| South | 3492855.68 | 146.68 |
| **Total** | **18667866.15** | **146.75** |
| **Lower Limb** |  |  |
| Midwest | 2310901.69 | 580.05 |
| Northeast | 10974837.34 | 460.49 |
| North | 1917519.35 | 455.03 |
| Southeast | 11192910.46 | 620.52 |
| South | 6243049.15 | 620.52 |
| **Total** | **32639218.00** | **542.81** |
| **Upper Limb** |  |  |
| Midwest | 307081.06 | 190.50 |
| Northeast | 947674.84 | 195.07 |
| North | 347324.63 | 182.32 |
| Southeast | 1289719.95 | 278.38 |
| South | 671213.18 | 259.15 |
| **Total** | **3563013.65** | **228.43** |

## DISCUSSION

Limb loss is a major problem both at individual and public health levels, resulting in decreased quality of life, working years and dependance. Traumatic amputation is one of its most common causes and an especially relevant one in developing countries, such as Brazil, where almost 75% of the population rely uniquely on governmental health care (11,14).

Traumatic amputations are generally caused by serious trauma, such as high-energy traffic accidents, explosions, and machine injuries (15). In these cases, limb salvage is often not possible. A study using data from the German Trauma Society showed that 13.6% of patients with traumatic amputations were able to undergo replantation, while 86.4% suffered limb loss (16). The decision whether or not to replant a limb depends on multiple factors, including the severity and extent of the injury, the presence of vascular lesions, the patient’s haemodynamic status and the resources available in the care service (17–19).

Between 2008 and 2023, 202,940 amputations were performed in the Brazilian public health system as a consequence of trauma. In terms of yearly incidence rate, an average of 9.058 per 100 000 inhabitants/year was observed compared to a total of 13.23 million globally found in 2019 (4). The figures remained stable during the observed period despite the previously reported tendency of a significant increase in overall lower limb amputations in the country (5).

For a continental-sized country such as Brazil, the incidence of traumatic amputations in the last 16 years was surprisingly even among most regions. While geographically and economically distinct, the south, the northeast, and the center-west demonstrated similar rates, varying from 122 per 100,000 inhabitants in the first to 116 in the latter. Perhaps even more interestingly, the southeast, home to São Paulo and Rio de Janeiro, the country’s two biggest cities (14), showed the lowest rate by far, with 77 amputations per 100,000 inhabitants. More studies will be necessary to clarify its reasons, but it may be explained by the more accessible health system in this region. The proportion of incidence of fingers or limb amputations also did not vary significantly across the country.

Regarding biological sex and age, the affected population had 4 times more men than women. Most were between 20 and 59 years, which aligns with previous reports (3–5,7,22). A possible explanation is partly occupational, with men often doing most of the high-risk physical and machinery work, in which accidents are more frequent. Young men also account for the majority of amputations caused by road traffic collisions. Together, those causes comprehend two of the most frequent etiologies leading to traumatic amputations (4).

These epidemiological results corroborate the impact of trauma on a country’s economic structure. Apart from the direct costs of hospitalization and rehab, the injury, which predominantly affects the most economically active population, culminates in absenteeism from work and loss in productive capacity.

Regarding the site, amputation was found to be most common in fingers, as shown in prior studies (5,7,22). This is probably due to the vulnerability of the hand to violent wounds and the lack of muscle protection. Finger wounds were particularly relevant in the pediatric age group, where they accounted for the vast majority of amputations. Traumatic amputation of the upper limbs occurred more rarely (7.7 percent), with an incidence similar to that described in developed countries (23).

Lower limb amputations accounted for a third of the sample, were increasingly frequent in the geriatric population, and differed less between the sexes than other types of amputations. A similar epidemiology was observed in a global, multicentre study, apparently because falls are the main mechanism of trauma in older adults (4). In addition, the presence of osteoporosis in this age group, which particularly affects women, can also potentiate the injury and lead to a more frequent need for amputation (4). Moreover, higher energy traumas are more likely to result in vascular injuries, carrying a worse prognosis (3,5,22). In the context of a rapidly aging country, the need for preventive measures adapted to different populations is evident.

The morbidity associated with leg lesions may help to explain why, in our study, victims of lower limb amputation required more intensive care admissions, longer hospital stays, and a higher lethality than the other patients. Regardless of advances in medical treatment, the overall lethality of 3,15, shown in our figures, remained relatively stable over the period studied. Nevertheless, the mortality rate is still lower than the 6% to 13% found in the literature (1,2).

As a universal public health system, the total cost of $54,870,097.79 spent in the period studied was covered in its totality by SUS, representing an expense of $34,29381 per year in traumatic amputations. Due to its severity, a patient with lower limb amputation was found to cost almost twice as much as the overall cost per patient ($542.81 to $270.38). It is also important to highlight that the sum does not include indirect hospital costs, rehab, or loss of working life years, which means that the total economic impact of this event for the country is probably higher.

This national epidemiological study, the first of its kind in Latin America, aims to present the main data on traumatic amputations in Brazil, including their impact on the national public health system. Similar to other types of trauma, identifying the patterns of these injuries is essential for their prevention. Therefore, our results can help guide strategies to reduce the significant impact and morbidity of traumatic amputations in our population.

Limitations of this study include those of population-based retrospective studies. In addition, as DATASUS is mainly based on epidemiological data from ICD-10 codes, the true number of traumatic amputations is likely to be underestimated due to coding errors. Finally, our analysis does not include private and supplementary healthcare systems, thus evaluating approximately three-fourths of the country’s population (11).

## CONCLUSION

Traumatic amputation in Brazil from 2008 to 2023 was most common among young men, affecting particularly the fingers but accounting for longer hospital stays and more ICU admissions when involving lower limbs. The mean duration of hospitalization was 4,57 days, culminating at a 3.15% lethality rate, which remained stable during the overlooked period. The total cost to the Brazilian public health system was $54,870,097.79, representing $34,29381 spent per year in traumatic amputations, with lower limb accounting for a figure twice as high per patient.

## Data Availability

All data produced in the present study are available upon reasonable request to the authors

https://datasus.saude.gov.br/

## APPENDIX

**Supplementary Table 1:** Traumatic Amputations per region of Brazil.

|  | Traumatic Amputations |  |  | p-value |
| --- | --- | --- | --- | --- |
|  | Fingers | Lower Limb | Upper Limb |  |
| Region |  |  |  |  |
| Midwest | 13350 (10.5) | 3984 (6.6) | 1612 (10.3) | <0.001 |
| Northeast | 36277 (28.5) | 23833 (39.6) | 4858 (31.1) |  |
| North | 11478 (9.0) | 4214 (7.0) | 1905 (12.2) |  |
| Southeast | 42294 (33.2) | 18038 (30.0) | 4633 (29.7) |  |
| South | 23813 (18.7) | 10061 (16.7) | 2590 (16.6) |  |
| Total | 127212 (100.0) | 60130 (100.0) | 15598 (100.0) |  |

**Supplementary Table 2:** Traumatic Amputations per age group.

|  | Traumatic Amputation |  |  | p-value |
| --- | --- | --- | --- | --- |
|  | Fingers | Lower Limb | Upper Limb |  |
| Age group (years) |  |  |  |  |
| 0 - 14 | 12494 (9.8) | 1055 (1.8) | 1294 (8.3) | <0.001 |
| 15 - 39 | 49427 (38.9) | 13154 (21.9) | 7292 (46.7) |  |
| 40 - 64 | 48860 (38.4) | 21750 (36.2) | 5673 (36.4) |  |
| 65 or older | 16431 (12.9) | 24171 (40.2) | 1339 (8.6) |  |
| Total | 127212 (100.0) | 60130 (100.0) | 15598 (100.0) |  |

**Table 3.** Amputations per age group.

**Supplementary Table 3:** Traumatic Amputations - days in hospital.

|  | Traumatic Amputations |  |  | p-value |
| --- | --- | --- | --- | --- |
|  | Fingers | Lower Limb | Upper Limb |  |
| Days in hospital |  |  |  |  |
| 0 - 2 | 86367 (67.9) | 10824 (18.0) | 8101 (51.9) | <0.001 |
| 3 - 7 | 30084 (23.6) | 28780 (47.9) | 5337 (34.2) |  |
| 8 - 14 | 7282 (5.7) | 12357 (20.6) | 1351 (8.7) |  |
| 15 or more | 3479 (2.7) | 8169 (13.6) | 809 (5.2) |  |
| Total | 127212 (100.0) | 60130 (100.0) | 15598 (100.0) |  |

**Supplementary Figure 1:**
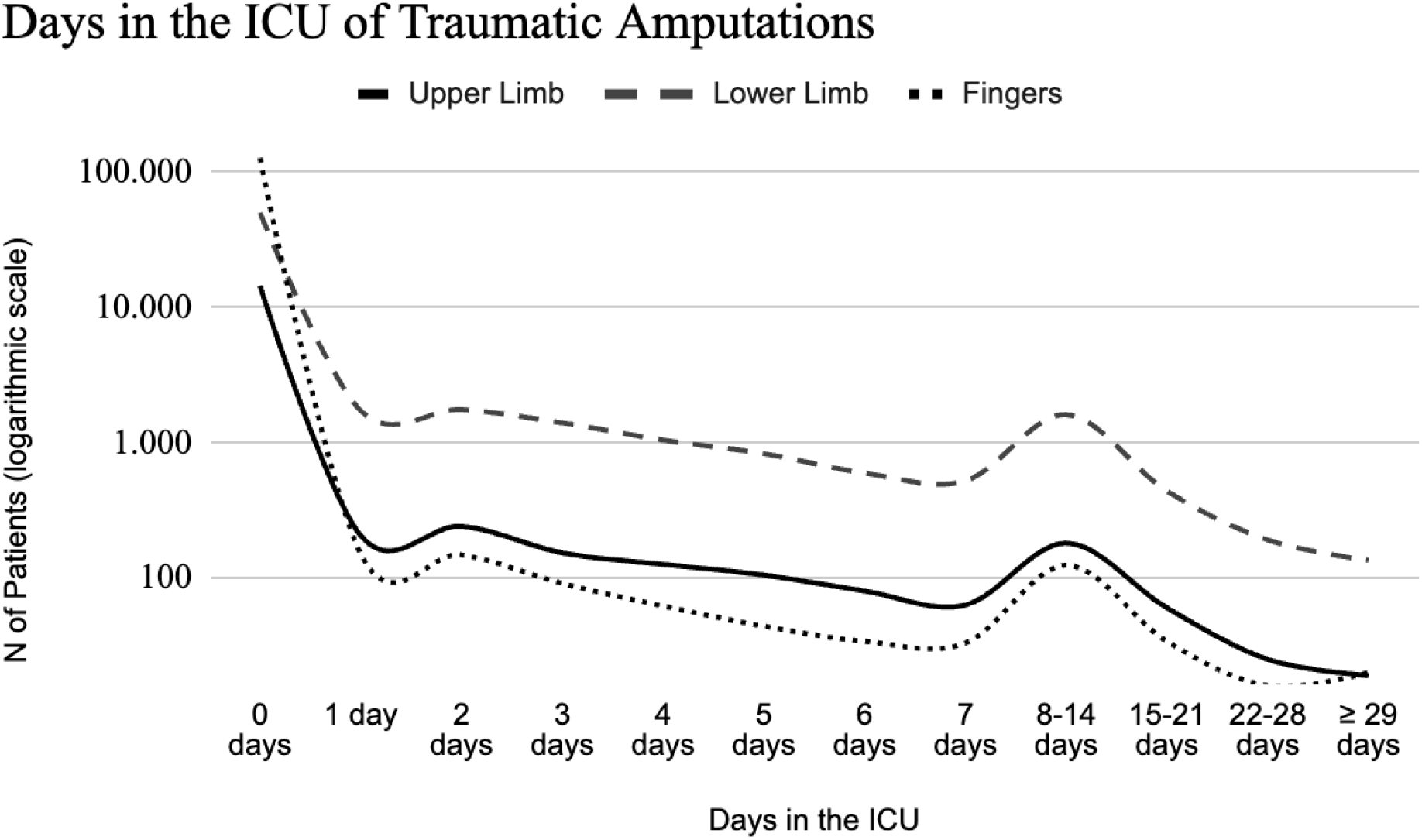
Traumatic Amputations - days in the intensive care unit (ICU).

